# Determinants and Incidence of Tuberculosis among adults living with HIV, who completed Isoniazid Preventive Therapy from 2019 to 2023, Mainland Tanzania

**DOI:** 10.64898/2026.06.10.26355305

**Authors:** Dosanto Felix Mlaponi, Veryeh Sambu, Joshua Stephen Mollel, Mgolegrace Smart Mabusi, Sanuni Ally Kessy, Jovin Tibenderana

## Abstract

**Objective:** This study aimed to determine TB incidence, median duration of Isoniazid Preventive Therapy protection (PLHIV), and factors associated with TB among adult People living with HIV (≥15 years) on ART who completed IPT

**Methodology:** This secondary analysis examined adult PLHIV who completed IPT using data from the CTC2 database (Jan 2019–Dec 2023, mainland Tanzania). Statistical analysis was conducted in STATA 15, with Cox regression used to identify factors associated with tuberculosis diagnosis. Proportional hazard assumptions were tested using Schoenfeld residuals scatter plots.

**Results:** A total of 38,407 adults living with HIV (≥15 years) were included in the analysis. The mean age was 36.7 years, with 63.0% female. The median follow-up time was 17.9 months, during which 901 incident TB cases were identified over 682,252.63 person-months, yielding an incidence rate of 1.40 per 1,000 person-months. The median time to TB development was 10.12 months. Female sex (aHR = 1.23; 95% CI: 0.86–1.75; p = 0.025), being unmarried (aHR = 1.20; 95% CI: 0.71–1.51; p = 0.034), and breastfeeding status (aHR = 1.12; 95% CI:1.02–1.56; p = 0.034) had higher risk of TB. Those aged 25–34 years (aHR = 0.55; 95% CI: 0.33–0.89; p = 0.015) and ≥45 years (aHR = 0.56; 95% CI: 0.32–0.95; p = 0.031) had a lower risk of TB. Geographically, those in Eastern (aHR = 0.21; 95% CI: 0.11–0.42; p < 0.001), Western (aHR = 0.37; 95% CI: 0.19–0.73; p = 0.004), and Lake zones (aHR = 0.78; 95% CI: 0.18–0.50; p < 0.001) had lower TB risk compared to the Northern zone. At Health centres (aHR = 0.57; 95% CI: 0.40–0.81; p = 0.002) and hospitals (aHR = 0.20; 95% CI: 0.11–0.39; p = 0.001) had reduced TB risk compared to dispensary

**Conclusion:** Among adults living with HIV on ART who completed IPT in mainland Tanzania, the incidence of TB remained low, confirming the protective benefit of IPT in reducing TB occurrence. Evidence shows IPT protection among PLHIV may last only about 10 months rather than the 18 months stated in the Tanzania HIV treatment guidelines, underscoring the need for earlier and intensified TB screening, revision of IPT guidelines, and consideration of alternative TB preventive therapy regimens.

## Introduction

People with HIV (PLHIV) are 20 to 37 times more likely to develop active tuberculosis (TB) from latent TB than those without HIV, making HIV infection the strongest risk factor for TB. According to the World Health Organization (WHO), isoniazid treatment reduces the likelihood of developing active tuberculosis (TB) by approximately 40% to 60% compared to the risk that would have been present without treatment [1]. This benefit is observed specifically among PLHIV.

Globally, the TB incidence rate (new cases per 100,000 population per year) rose by 3.6% between 2020 and 2021, reversing declines of about 2% per year for most of the previous 2 decades. The global rise in the TB incidence rate may be largely attributed to the COVID-19 pandemic, which disrupted healthcare services, reduced access to care, and diverted resources away from TB program[2]. Additionally, infection control challenges in healthcare settings and the fear and stigma associated with respiratory symptoms led to fewer people seeking medical care for TB, reversing the previous trend of declining TB incidence rates observed over the past two decades[2]

The burden of the disease is especially high in Sub-Saharan Africa (SSA), which contributes more than 25% of global tuberculosis (TB) cases and records the highest rates of TB/HIV co-infection. Tanzania is among the 30 high TB burden countries, with an estimated incidence of 183 cases per 100,000 population in 2023[3], [4]. TB case notification rate has generally been increasing since 2015, with a 16% rise recorded between 2015 and 2023[4]. While HIV testing for TB patients rose from 88% to 99%, the proportion of HIV-positive TB cases fell from 36% to 24% in 2019[5]

According to the Tanzania health sector strategic plan IV, the targeted plan was to reduce the TB incidence to lower than 231 cases per 100,000 by 2020, but the target was not achieved, and it only attained the incidence of 273 cases per 100,000, which was higher than expected This means that the planned reduction in TB incidence was not achieved and that the incidence rate was higher than the targeted goal[4], [5]. The current health sector strategic plan V (2021-2026) has the aim to reduce the incidence of TB, aiming to accelerate the reduction of the tuberculosis (TB) incidence rate, with specific targets to reduce it by 27% compared to 2015 levels[6]. The current study aimed to determine TB incidence, the median duration of IPT protection, and factors associated with TB among adult PLHIV (≥15 years) on ART who completed IPT (2015–2023) in mainland Tanzania.

This study will provide critical evidence to inform policy decisions regarding both the current incidence of tuberculosis (TB) among people living with HIV (PLHIV) and the median duration of protection conferred by isoniazid preventive therapy (IPT). By generating robust, context-specific data, it aims to guide optimization of IPT implementation strategies, including timing, duration, and targeting of therapy, ultimately strengthening TB prevention efforts and improving health outcomes within this high-risk population.

## Material and methods

### Study design and setting

This was a retrospective cohort design which utilized secondary data from the CTC2 database of adult PLHIV who completed IPT use and collected over five years from 1^st^ January, 2019 to 31^st^ December, 2023. Each patient was assigned a unique medical registration code, and relevant clinical and demographic data were systematically extracted from follow-up records documented in the hospital’s medical files. Despite the extensive data collection period, the observation time for each participant was limited to 18 months after the completion of IPT. This 18-month follow-up period was chosen to ensure a consistent and focused assessment of the therapy’s effectiveness and any subsequent health outcomes for the participants.

The secondary analysis of routinely collected data was conducted across all 26 regions of the mainland, covering a total population of 61.7 million people, with 50.4% being female and 49.6% male. Of this population, about 29.3 million live in urban areas, while around 32.4 million reside in rural areas [7]. In 2022, the estimated number of adults living with HIV (PLHIV) in Tanzania was approximately 1,548,000, and about 7,805 facilities were providing HIV care and treatment on the mainland [8]

### Study population

All PLHIV patients aged 15 years and above living in mainland Tanzania who completed IPT between 1^st^ January 2019 and 31^st^ June 2023.

### Data Source

The data were extracted from the CTC2 database at NACP, combining all patients’ records attending CTC. Standardized patient management forms, known as CTC2 cards, are used in health facilities to record patient information. These cards capture key details including socio-demographic characteristics, WHO clinical stage, weight, CD4 count, ART regimen, date of ART initiation, tuberculosis screening results, nutritional status, height, ART adherence, mortality information, and haemoglobin levels. Within each CTC clinic, data are transferred to the electronic CTC2 database for storage and future use when needed. Each patient is identified using a unique identification number assigned at the time of enrolment

### Eligibility criteria

All PLHIV on ART aged 15 years and above who completed IPT between 2019 and 2023 were included in the study, while PLHIV aged 15 years and above who have been treated for TB during IPT use were excluded.

### Power of the study

The power of the study was calculated based on the formula for two proportions according to Kirkwood et al (2018), considering the total number of TB cases, which was 901. The formula to calculate the power is given below;

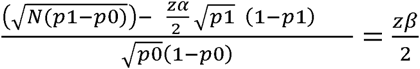

Where; N: Number of eligible adults PLWHIV who completed IPT and were diagnosed with TB was 901 from 2019 to 2023. p1: Estimated Proportion of PLHIV with diagnosis TB obtained in this study was 2.35 %. p0: Proportion of TB under null hypothesis=5.8% [9], Zα/2: Standard normal value for 5% precision (1.96), Zβ/2: Standard normal value for power. Z score: calculation from the formula above 2.4, which corresponds to 0.99180 in the z score table, this gives the statistical power of this study, 99.0 %

### Study Variables

The main outcomes in this study were the occurrence of TB among people living with HIV (PLHIV) on antiretroviral therapy (ART), analyzed as a time-to-event binary outcome. The independent variables were age, sex, marital status, baseline CD4 cell count, baseline WHO HIV clinical stage were grouped into stage I, II, III, and IV, baseline functional status (working, ambulatory and Bedridden), Baseline BMI category, Year of ART start, health facility ownership, health facility level, Pregnancy status, ART adherence which was documented as poor and good based on the pill count and Viral suppression and baseline viral load.

Nutrition status was categorized into four groups according to BMI scales. Participants with a BMI of <18.5 kg/m^2^ (Underweight), 18.5–24.9kg/m^2^ (Normal weight), 25.0– 29.9kg/m^2^ (Overweight), and ≥30.0 kg/m^2^ (Obesity). ART regimens were categorized as first-line and second-line based on the Tanzania HIV guideline

### Data management plan

The letter requesting permission to use the CTC2 database was sent by KCMC University to the permanent secretary, Ministry of Health, in the United Republic of Tanzania. Permission to use the National HIV care and treatment Data (CTC2) for research and publication was obtained. The Do file was sent to the head of the strategic information unit at the National Aids, STIs, and Hepatitis Control program (NASHCoP), and finally, the data for this research was successfully extracted on 15^th^ June 2024. After extraction, the data were evaluated for quality, completeness, and relevance to ensure reliability and validity of the data before analysis. The median and mean imputation methods were used for missing data. Some of the variables were omitted during data cleaning.

### Statistical analysis

Data analysis was performed using STATA version 15.0 Special edition (Stata Corp, College Station, TX) software. Before analysis, data were explored, examined, cleaned, and checked for missing values and completeness. Descriptive analysis was done by summarizing data using means with standard Deviation (SD) and Median (IQR), depending on the level of skewness of continuous variables; frequencies and proportions were used for categorical variables. Kaplan Meier cumulative probability curves were used to determine and compute the median time till developing TB among PLHIV on ART who completed IPT. Associations between categorical variables were assessed using chi-square tests and Fisher’s exact tests with the level of significance set at P-value ≤ 0.05.

The patient’s time at risk for TB was defined as the time from when the client completed IPT until 18 months of follow-up, where each client was followed only for 18 months. Those who completed the follow-up period without developing TB, those who died during the follow-up period with other causes of death and not death from TB, and those who were lost to follow-up during the study were termed as censored observations, which means their person-months contributed were considered during analysis. The overall TB incidence rate was determined as the total number of TB cases per total person-months recorded. A multivariable Cox proportional hazards regression model was used to obtain factors associated with diagnosis of TB after accounting, and the Hazard ratio for factors that were significantly associated with diagnosis of TB was reported. A level of significance was set at P-value ≤ 0.05.

### Ethical considerations

The secondary data analysis obtained ethical approval from the Institutional Review Board (IRB) called KCMC University Research Ethics Review Committee (CRERC) (Approval ID: PG128/2023, dated 27th September, 2023). Permission to use the CTC2 database for the analyses was obtained from the Permanent Secretary, Tanzania Ministry of Health, and the National AIDS, STIs and Hepatitis Control Program (NASHCoP). Informed consent was not required because patients’ records were anonymized before access

## RESULTS

Among a total of 1,529,063 PLHIV found in the dataset from January 2019 to December 2023, a total of 1,490,656 who were aged <15 years and those with no complete data during the study period were excluded. Finally, 38,407 PLHIV (With or without TB diagnosis) were eligible for this study based on the inclusion criteria **Fig 1**.

**Fig 1:**
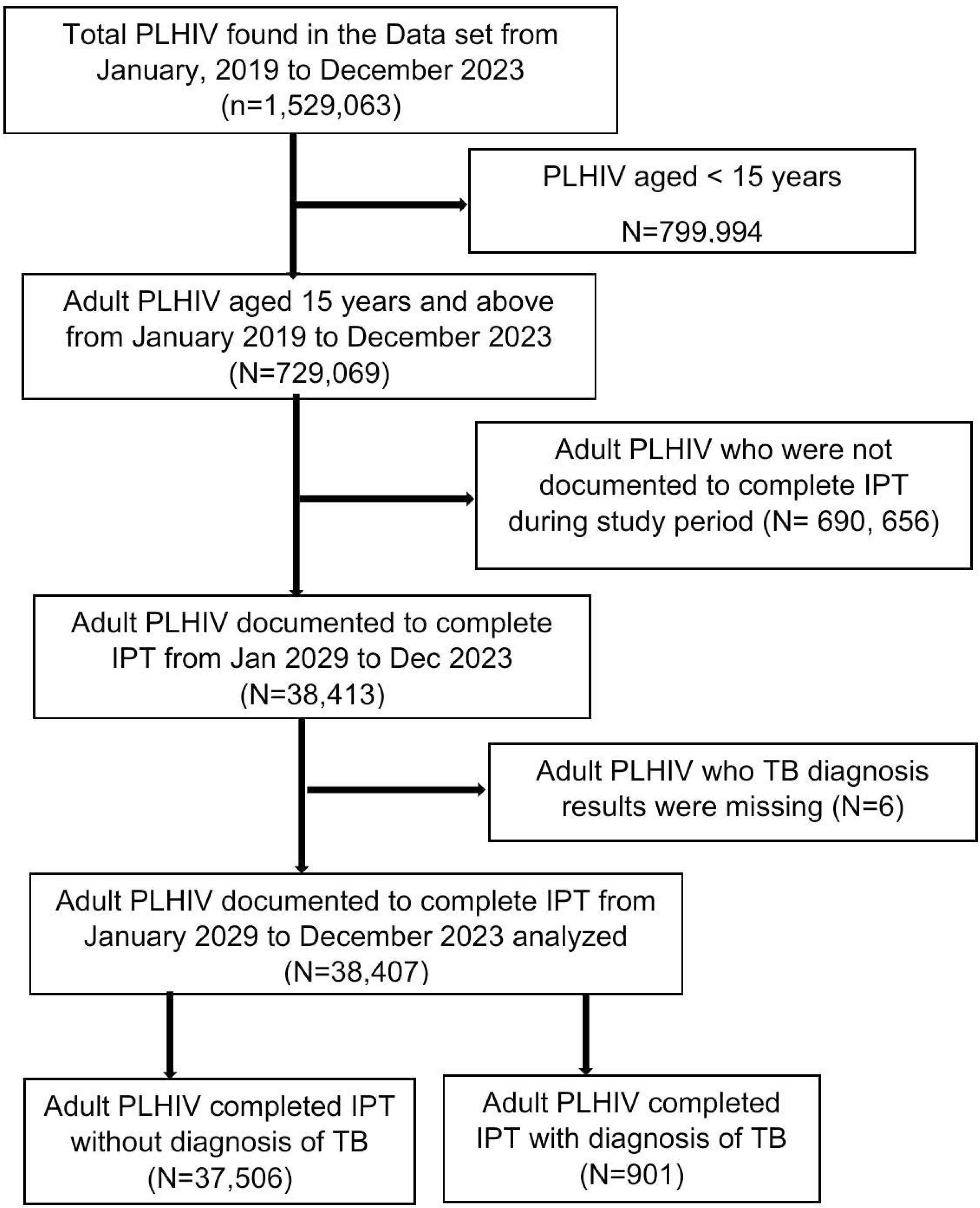
The flow chart of recruitment of participants from the CTC2 data set of People living with HIV(PLHIV) in Mainland Tanzania Among 1,529,063 People living with HIV(PLHIV) found in the CTC2 data set from January 2019 to December 2023, a total of 38,407 PLHIV with and without TB at diagnosis who completed Isoniazid preventive therapy (IPT) were eligible for this study.

### Social demographic characteristics

Among 38,407 PLHIV enrolled, the mean (SD) age was 36.7± (11.3) years. The largest group was 25-34 years old, 12,597 (32.80%). The majority of participants were female, 23,365 (60.84%), and 25,372 (66.06%) of individuals were married. Geographically, the Lake zone had the highest population representation with 14,407 (37.51%) followed by the Eastern zone, 6,403(16.67%) see **Table 1**.

**Table 1:** Social demographic characteristics of adult PLHIV aged 15 years and above on ART after IPT completion from Jan 2019 to June 2023 (N= 38,407)

| Variable | Frequency (N) | Percentage (%) |
| --- | --- | --- |
| **Age (years)\*** |  |  |
| 15-24 | 4,632 | 12.06 |
| 25-34 | 12,597 | 32.80 |
| 35-44 | 11,125 | 28.97 |
| ≥ 45 | 8,715 | 22.69 |
| Missings | 1338 | 3.48 |
| Mean age (SD) | 36.7± (11.3) |  |
| **Sex\*** |  |  |
| Male | 13,704 | 35.68 |
| Female | 23,365 | 60.84 |
| Missings | 1,338 | 3.48 |
| **Marital status\*** |  |  |
| Married | 25,372 | 66.06 |
| Unmarried | 11,542 | 30.05 |
| Missings | 1,493 | 3.89 |
| **Zones\*** |  |  |
| Northern | 2,885 | 7.51 |
| Central | 1,752 | 4.56 |
| Eastern | 6,403 | 16.67 |
| Southern | 1,059 | 2.76 |
| Western | 3,505 | 9.13 |
| Southern highlands | 3,501 | 9.12 |
| Southwest highlands | 3,327 | 8.66 |
| Lake | 14,407 | 37.51 |
| Missings | 1,568 | 4.08 |

Caption: \* Variables with the missing values in the CTC2 dataset.

### Clinical characteristics

The majority of individuals 36,381 (98.72%) had good adherence to antiretroviral therapy (ART)which was measured by pill count, clinical attendance and viral suppression. In terms of the WHO clinical stage, most individuals 22,465 (58.49%) were in stage I. Also, majority of PLHIV 32,687 (88.11%) had normal BMI. Nearly all PLHIV 38,387 (99.95%) were working. A significant portion of the PLHIV 14,939 (38.90%) were pregnant. Regarding the type of health facility attended, most 17,777(46.29%) went to dispensaries The majority of PLHIV 22,748 (59.35%) were receiving care from public health facilities (59.23%). Only few PLHIV (5.02%) were breastfeeding. Most of PLHIV 114 (52.29%) had a CD4 count greater than 350 cells/mm³ with a median of 400 (IQR 11.94-1637). Most participants 1,770 (84.69%) had viral load of less than 1,000 copies/mL with a median viral load of 47 (IQR 21-473000). Regarding the year that ART was started, 14,648 (38.14%) of PLHIV began in 2019, followed by 12,721 (33.12%) in 2020, 5,773 (15.03%) in 2021, 4,524 (11.78%) in 2022, and 741 (1.93%) in 2023. This shows a higher initiation rate in earlier years, with a decline in more recent years, see in **Table 2**.

**Table 2:**
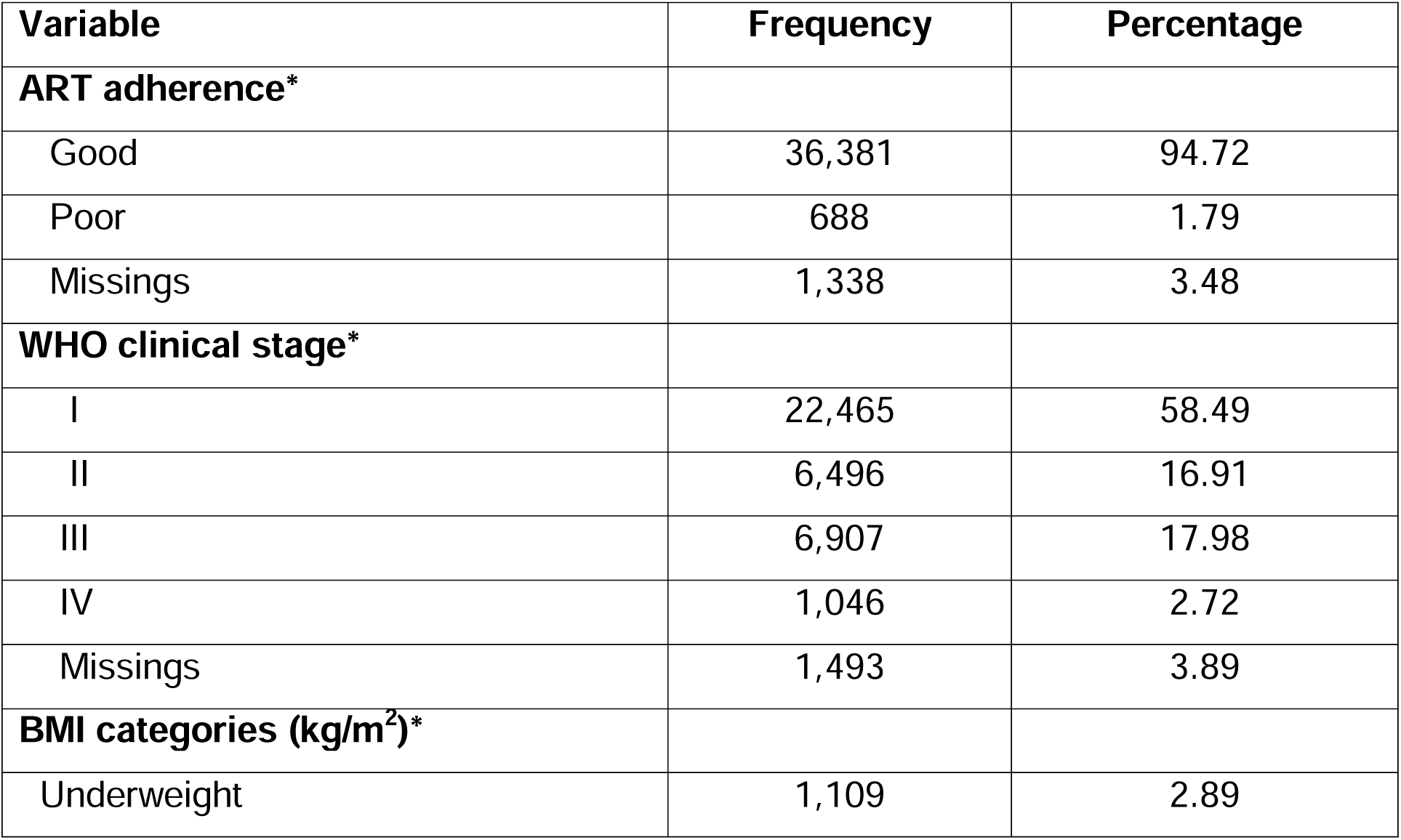
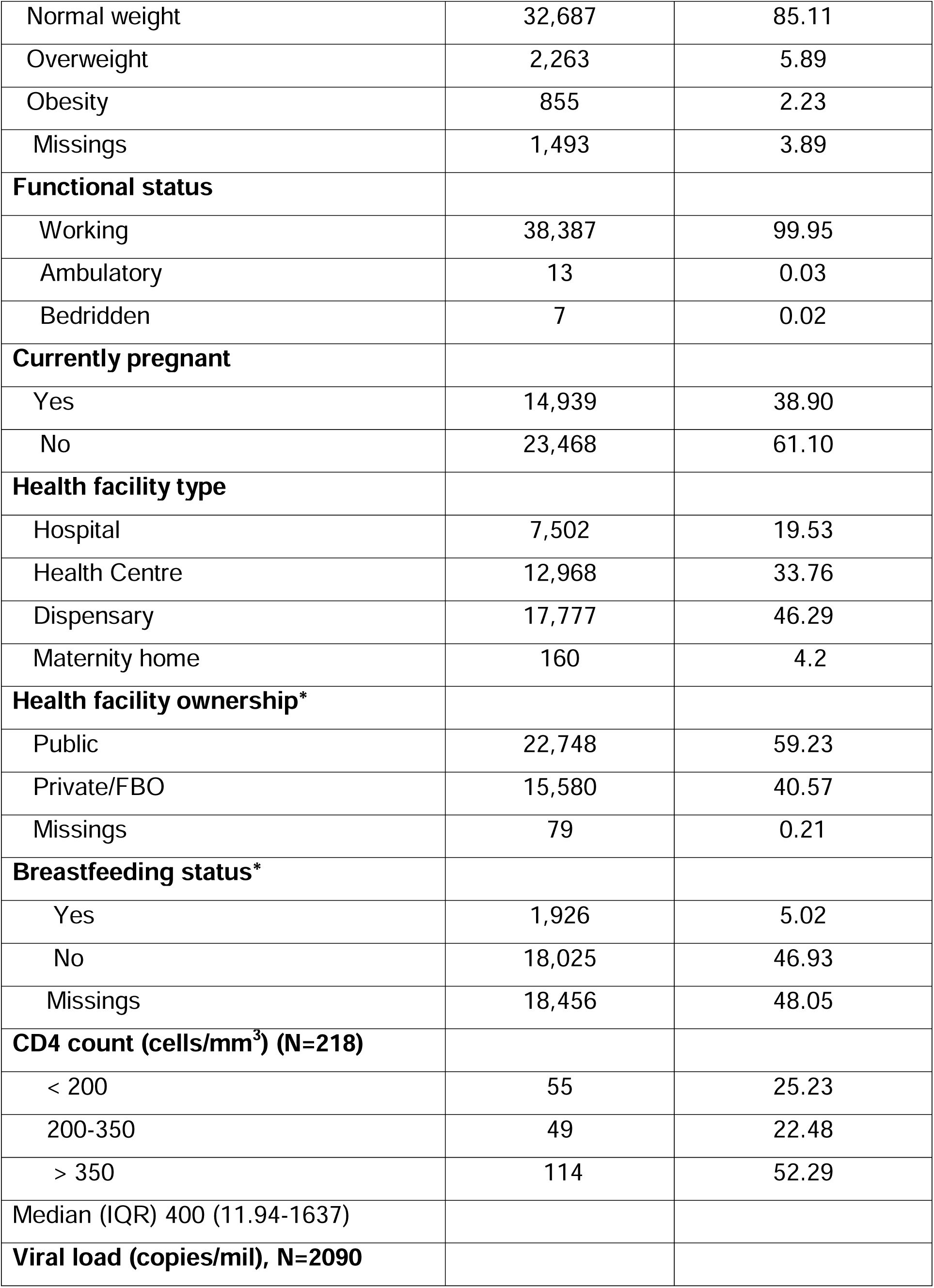
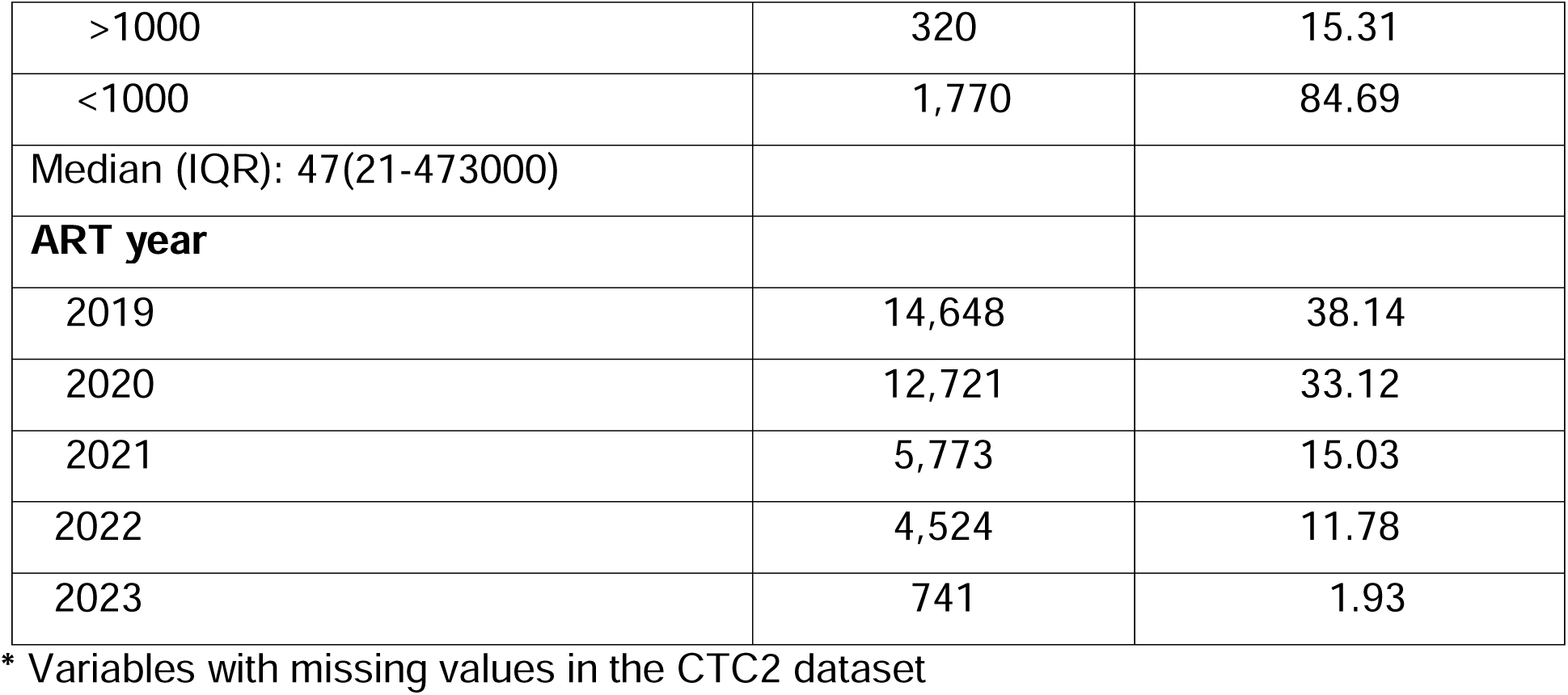
Clinical characteristics of adult PLHIV aged 15 years and above current on ART completed IPT in Tanzania mainland from January 2019 to June 2023 (N= 38,407)

| Variable | Frequency | Percentage |
| --- | --- | --- |
| **ART adherence\*** |  |  |
| Good | 36,381 | 94.72 |
| Poor | 688 | 1.79 |
| Missings | 1,338 | 3.48 |
| **WHO clinical stage\*** |  |  |
| I | 22,465 | 58.49 |
| II | 6,496 | 16.91 |
| III | 6,907 | 17.98 |
| IV | 1,046 | 2.72 |
| Missings | 1,493 | 3.89 |
| **BMI categories (kg/m<sup>2</sup>)\*** |  |  |
| Underweight | 1,109 | 2.89 |
| Normal weight | 32,687 | 85.11 |
| Overweight | 2,263 | 5.89 |
| Obesity | 855 | 2.23 |
| Missings | 1,493 | 3.89 |
| **Functional status** |  |  |
| Working | 38,387 | 99.95 |
| Ambulatory | 13 | 0.03 |
| Bedridden | 7 | 0.02 |
| **Currently pregnant** |  |  |
| Yes | 14,939 | 38.90 |
| No | 23,468 | 61.10 |
| **Health facility type** |  |  |
| Hospital | 7,502 | 19.53 |
| Health Centre | 12,968 | 33.76 |
| Dispensary | 17,777 | 46.29 |
| Maternity home | 160 | 4.2 |
| **Health facility ownership\*** |  |  |
| Public | 22,748 | 59.23 |
| Private/FBO | 15,580 | 40.57 |
| Missings | 79 | 0.21 |
| **Breastfeeding status\*** |  |  |
| Yes | 1,926 | 5.02 |
| No | 18,025 | 46.93 |
| Missings | 18,456 | 48.05 |
| **CD4 count (cells/mm<sup>3</sup>) (N=218)** |  |  |
| < 200 | 55 | 25.23 |
| 200-350 | 49 | 22.48 |
| > 350 | 114 | 52.29 |
| Median (IQR) 400 (11.94-1637) |  |  |
| **Viral load (copies/ml), N=2090** |  |  |
| >1000 | 320 | 15.31 |
| <1000 | 1,770 | 84.69 |
| Median (IQR): 47(21-473000) |  |  |
| **ART year** |  |  |
| 2019 | 14,648 | 38.14 |
| 2020 | 12,721 | 33.12 |
| 2021 | 5,773 | 15.03 |
| 2022 | 4,524 | 11.78 |
| 2023 | 741 | 1.93 |

\* Variables with missing values in the CTC2 dataset

### Incidence of TB diagnosis

Among 901 PLHIV diagnosed with TB, females exhibited a higher rate of TB at 1.44 per 1000 person-months compared to male 1.27 per 1000 person-months. PLHIV participants aged 15-24 years had higher incidence rate of 1.49 per 1000 person-months compared to those above 25 years. Unmarried individuals have a higher rate of health events at 1.45 per 1000 person-months compared to married individuals. The Northern and Central zones report the highest rates at 2.73 and 2.71 per 1000 person-months, respectively, while the Eastern zone has a relatively lower rate of 1.04 per 1000 person-months. From the CTC2 data set the findings show that most individuals (99.9%) are working, indicating good functional health status overall. Individuals with viral loads <1000 copies/mL had a higher rate of diagnosis of TB at 1.22 per 1000 person-months compared to those with >1000 copies/mL at 0.72 per 1000 person-months. The rate is significantly lower for individuals with CD4 counts <200 cells/mm³ at 0, whereas those with counts >200 cells/mm³ had a rate of 0.72 per 1000 person-months. Underweight, overweight, and obese individuals have higher rates of health events (1.92, 1.86, and 1.90 per 1000 person-months, respectively) compared to those with normal weight (1.31 per 1000 person-months). Pregnant individuals have a slightly lower rate at 1.29 per 1000 person-months compared to non-pregnant individuals at 1.43 per 1000 person-months. PLHIV in attending dispensaries and health centres had the higher rate of health events at 1.86 per 1000 person-months and 1.26 per person-months, respectively, compared to those attending the hospital, 0.42(0.32, 0.54). Adult PLHIV who attended Public health facilities had a higher rate of diagnosis of TB, 1.99 per 1000 person-months, compared to those who attended private/FBO facilities (0.47 per 1000 person-months), see in **table 3**

**Table 3:** Incidence of TB among adult PLHIV aged 15 years and above, current on ART after completion of IPT in Tanzania mainland from January 2019 to June 2023 (N= 901) Legend: FBO-Faith Based Organisation; BMI-Body Mass Index, *Sample size for Viral load, (N=152) and CD4 count is (N= 218), # Variable with the missing value in CT2 dataset.

| Variable | Number of cases | Rate/1000(95%CI) | Person-Month |
| --- | --- | --- | --- |
| **Sex** |  |  |  |
| Male | 307 | 1.27(1.13, 1.42) | 242564.39 |
| Female | 594 | 1.44(1.33 1.56) | 413172.31 |
| **Age in years** |  |  |  |
| 15-24 | 121 | 1.49(1.25, 1.78) | 81230.78 |
| 25-34 | 303 | 1.36(1.22, 1.52) | 222762.48 |
| 35-44 | 264 | 1.34(1.19, 1.51) | 197141.75 |
| ≥45 | 213 | 1.38(1.20, 1.58) | 154601.68 |
| **Marital status** |  |  |  |
| Married | 603 | 1.34(1.23, 1.45) | 450770.86 |
| Unmarried | 298 | 1.45(1.30, 1.63) | 204965.83 |
| **Zones** |  |  |  |
| Northern | 139 | 2.73(2.31, 3.23) | 50847.57 |
| Central | 84 | 2.71(2.19, 3.35) | 31027.76 |
| Eastern | 94 | 0.82(0.67, 1.01) | 114227.99 |
| Southern | 43 | 2.28(1.69, 3.07) | 18870.89 |
| Western | 81 | 1.29(1.04, 1.61) | 62467.31 |
| Southern highlands | 120 | 1.93(1.62, 2.31) | 62127.13 |
| Southwest highlands | 73 | 1.23(0.98, 1.55) | 59208.77 |
| Lake | 267 | 1.04(0.93, 1.18) | 255616.46 |
| **Viral load, (N=152)\*** |  |  |  |
| <1000 | 149 | 1.22(0.23, 2.23) | 122177.40 |
| >1000 | 3 | 0.72(1.04, 1.43) | 4171.65 |
| **CD4 count, (N= 218)\*** |  |  |  |
| <200 | 0 | 0 | 0 |
| >200 | 3 | 0.72(0.23, 2.22) | 4189.55 |
| **WHO clinical stage** |  |  |  |
| I | 556 | 1.39(1.28, 1.51) | 398974.38 |
| II | 159 | 1.39(1.18, 1.61) | 115320.14 |
| III | 171 | 1.39(1.20, 1.62) | 122786.14 |
| IV | 15 | 0.80(0.48, 1.33) | 18656.045 |
| **BMI** |  |  |  |
| Underweight | 38 | 1.92(1.40, 2.63) | 19784.001 |
| normal weight | 759 | 1.31(1.22, 1.40) | 580306.83 |
| Overweight | 75 | 1.86(1.48, 2.33) | 40391.59 |
| obesity | 29 | 1.90(1.32, 2.74) | 15254.27 |
| **Pregnancy status** |  |  |  |
| No | 574 | 1.43(1.31, 1.55) | 402591.00 |
| Yes | 327 | 1.29(1.159, 1.43) | 253145.70 |

| Level of health facility |  |  |  |
| --- | --- | --- | --- |
| Dispensary | 570 | 1.86(1.71, 2.02) | 306356.60 |
| Health Centre | 277 | 1.26(1.12, 1.42) | 219718.79 |
| Hospital | 54 | 0.42(0.32, 0.54) | 128318.50 |
| Facility ownership |  |  |  |
| Private/FBO | 125 | 0.47(0.39, 0.56) | 265624.84 |
| Public | 776 | 1.99(1.86, 2.14) | 388769.05 |
| Breastfeeding status# |  |  |  |
| No | 461 | 1.49(1.36, 1.64) | 308523.00 |
| Yes | 69 | 2.01(1.59, 2.54) | 34357.92 |
| Missings | 380 | - | - |

\* Variables with missing values in the CTC2 dataset

#### (a) Median disease-free survival time to diagnosis of TB

The overall median disease-free survival time to diagnosis of TB among adult PLHIV on ART after completion of IPT ranged from 5.8 to 24.6 months, with the median survival time of 10.12 months of censoring as seen in the Kaplan Meier survival graph, and the curve has several steep declines in the early to mid-analysis time, see **Fig 2**.

**Fig 2.**
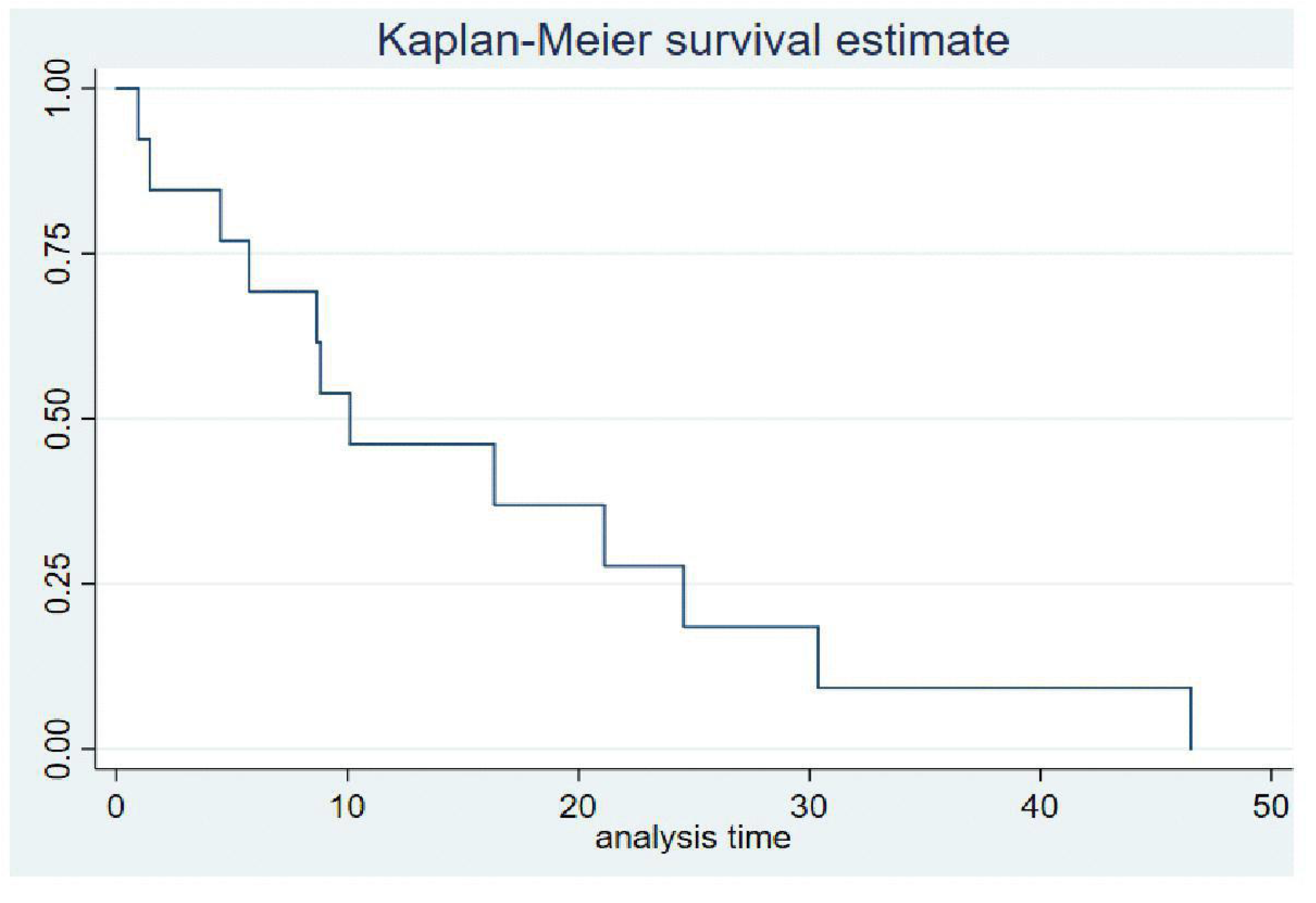
Kaplan –Meier survival estimate for median duration of developing TB among adult PLHIV aged 15 years and above after completing IPT.

#### (b) Median survival time to diagnosis of TB

After stratifying the median survival time to diagnosis of TB by male sex and female sex, the survival time to diagnosis of TB among male PLHIV after completing IPT ranged from 0.95 to 21.1 months, with a median survival time of 10.10. This was higher compared to female PLHIV, whose survival time ranged from 5.75 to 24.57 months, with a median survival time of 8.83 months, see **Fig 3**.

**Fig 3.**
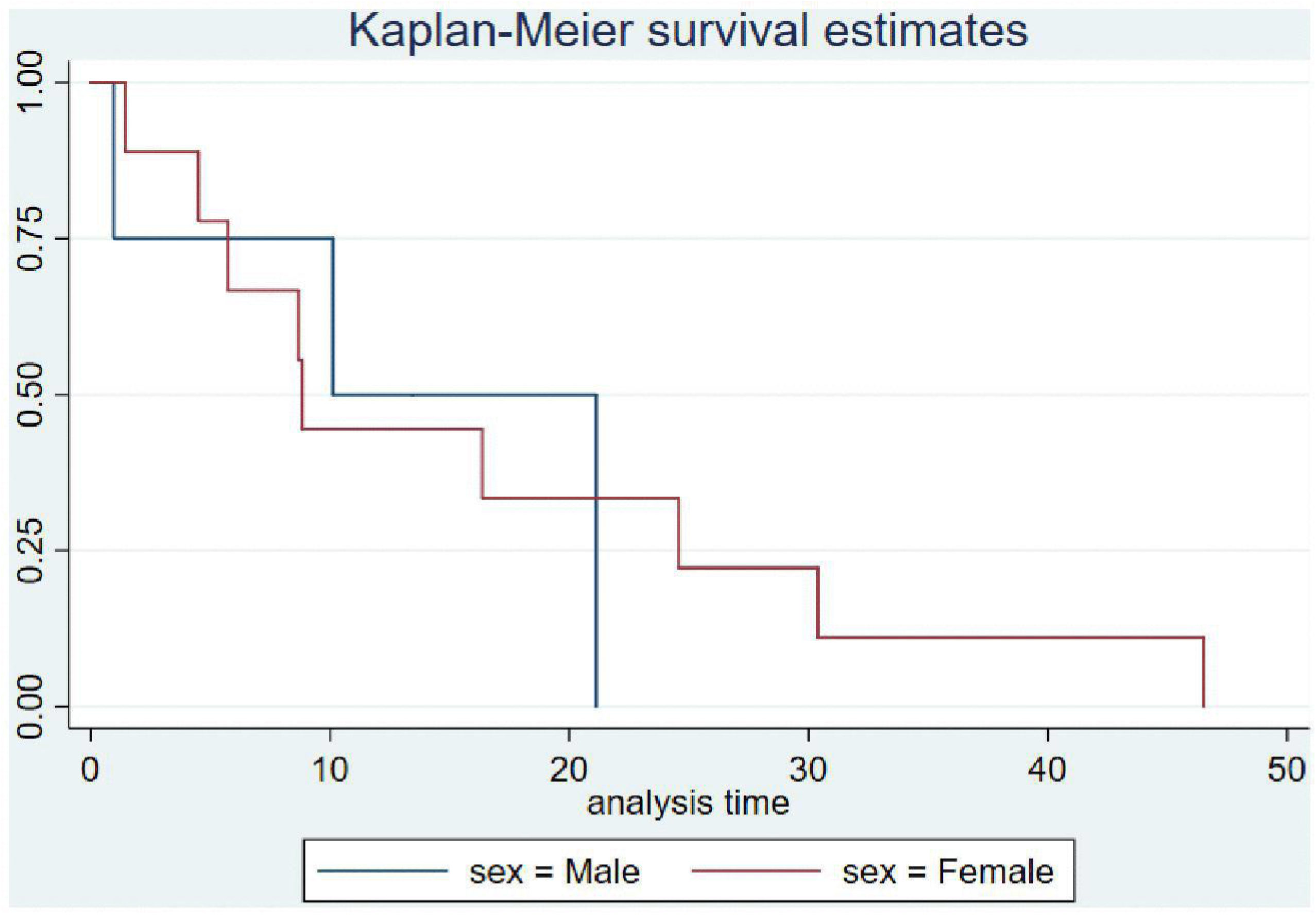
Kaplan –Meier survival estimate for median duration of developing TB among adult PLHIV aged 15 years and above after completing IPT.

### Factors associated with the diagnosis of Tuberculosis

In the univariate Cox proportional hazards analysis, several factors were associated with the hazard of tuberculosis (TB) diagnosis among people living with HIV (PLHIV). Unmarried individuals had a higher hazard of TB compared to married individuals (HR = 1.19; 95% CI: 1.03–1.38; *p* = 0.019). With respect to geographical zones, compared to Northern zone there was a significantly lower hazards of TB observed in the Eastern (HR = 0.30; 95% CI: 0.23–0.39; *p* < 0.001), Western (HR = 0.47; 95% CI: 0.36–0.62; *p* < 0.001), Southern Highlands (HR = 0.71; 95% CI: 0.55–0.90; *p* < 0.005), Southwest Highlands (HR = 0.45; 95% CI: 0.34–0.59; *p* < 0.001), and Lake zones (HR = 0.38; 95% CI: 0.31–0.47; *p* < 0.001). Individuals attending public health facilities had a markedly higher hazard of TB compared to those attending private or faith-based organization (FBO) facilities (HR = 4.24; 95% CI: 3.51–5.12; *p* < 0.001). In terms of disease stage, participants in WHO Stage IV had a higher hazard of TB compared to those in Stage I (HR = 1.28; 95% CI: 1.02–1.76; *p* = 0.036). Similarly, breastfeeding individuals had a higher hazard of TB compared to non-breastfeeding individuals (HR = 1.34; 95% CI: 1.04–1.73; *p* = 0.022). Poor adherence to treatment was also associated with an increased hazard of TB (HR = 1.42; 95% CI: 1.03–1.85; *p* = 0.021). Additionally, compared to dispensaries, attending health centres (HR = 0.68; 95% CI: 0.59–0.78; *p* < 0.001) and hospitals (HR = 0.23; 95% CI: 0.17–0.29; *p* < 0.001) was associated with a significantly lower hazard of TB, see **Table 4**.

**Table 4:** Cox proportional hazard analysis of factors associated with Tuberculosis incidence among adult PLHIV aged 15 years and above after completion of IPT (N=901)

| Variable | CHR (95%CI) | P VALUE | AHR (95% CI) | P VALUE |
| --- | --- | --- | --- | --- |
| **Sex** |  |  |  |  |
| Male | 1 |  | 1 |  |
| Female | 1.14(0.98, 1.30) | 0.070 | 1.23 (0.86, 1.75) | 0.025 |
| **Age in years** |  |  |  |  |
| 15-24 | 1 |  | 1 |  |
| 25-34 | 0.91(0.74, 1.13) | 0.493 | 0.55(0.33,0.89) | 0.015 |
| 35-44 | 0.89 (0.72, 1.11) | 0.332 | 0.65 (0.40, 1.06) | 0.086 |
| ≥45 | 0.92(0.74, 1.16) | 0.493 | 0.56(0.32, 0.95) | 0.031 |
| **Marital status** |  |  |  |  |
| Married | 1 |  | 1 |  |
| Unmarried | 1.19(1.03, 1.38) | 0.019 | 1.20(0.71, 1.51) | 0.0345 |
| **Zones** |  |  |  |  |
| Northern | 1 |  |  |  |
| Central | 0.99(0.76, 1.29) | 0.944 | 0.76(0.53,2.34) | 0.564 |
| Eastern | 0.30(0.23, 0.39) | <0.001 | 0.21(0.11, 0.42) | <0.001 |
| Southern | 0.83(0.59, 1.17) | 0.297 | 0.71(0.27, 1.89) | 0.495 |
| Western | 0.47(0.36,0.62) | <0.001 | 0.37(0.19, 0.73) | 0.004 |
| Southern highlands | 0.71(0.55, 0.90) | <0.005 | 0.56(0.29, 1.11) | 0.098 |
| Southwest highlands | 0.45(0.34, 0.59) | <0.001 | 0.56(0.29, 1.08) | 0.084 |
| Lake | 0.38(0.31,0.47) | <0.001 | 0.78(0.18, 0.50) | <0.001 |
| **BMI** |  |  |  |  |
| Normal weight | 1 |  |  |  |
| Underweight | 1.64(1.02,1.74) | <0.001 | 1.06(0.70, 1.61) | 0.052 |
| Overweight | 1.01(0.78, 1.31) | 0.915 | 1.23(0.56,1.45) | 0.723 |
| Obesity | 1.04(0.71, 1.52) | 0.848 | 1.35(0.95,2.56) | 0.534 |
| **Viral load** |  |  |  |  |
| >1000 | 1 |  | 1 |  |
| <1000 | 1.69(0.54, 5.32) | 0.365 | 1.93(0.61, 6.07) | 0.264 |
| **Health facility ownership** |  |  |  |  |
| Private/FBO | 1 |  | 1 |  |
| Public | 4.24(3.51, 5.12) | <0.001 | 3.92(2.44, 6.28) | 0.001 |
| **WHO clinical stage** |  |  |  |  |
| I | 1 |  | 1 |  |
| II | 1.12(0.83, 1.18) | 0.905 | 1.54(0.74,2.35) | 0.756 |
| III | 0.99(0.84, 1.19) | 0.994 | 0.76(0.17,1.29) | 0.624 |
| IV | 1.28(1.02,1.76) | 0.036 | 1.01(1.04, 2.31) | 0.042 |
| **Level of health facility** |  |  |  |  |
| Dispensary | 1 |  | 1 |  |
| Health Centre | 0.68(0.59, 0.78) | <0.001 | 0.57(0.40,0.81) | 0.002 |
| hospital | 0.23(0.17, 0.29) | <0.001 | 0.20(0.11, 0.39) | 0.001 |
| **Breastfeeding** |  |  |  |  |
| No | 1 |  | 1 |  |
| Yes | 1.34(1.04, 1.73) | 0.022 | 1.12(1.02,1.56) | <0.0345 |
| **ART Adherence** |  |  |  |  |
| Good | 1 |  | 1 |  |
| Poor | 1.42(1.03, 1.85) | 0.021 | 1.31 (0.05, 2.45) | <0.043 |

**\*Statistically significant, p<0.05.**

In the multivariable (adjusted) Cox proportional hazards analysis, several factors remained significantly associated with TB diagnosis. Female participants had a higher hazard of TB compared to males (aHR = 1.23; 95% CI: 0.86–1.75; *p* = 0.025). Participants aged 25–34 years and ≥45 years had a significantly lower hazard of TB compared to those aged 15–24 years (aHR = 0.55; 95% CI: 0.33–0.89; *p* = 0.015) and (aHR = 0.56; 95% CI: 0.32–0.95; *p* = 0.031) respectively. Unmarried individuals continued to have a higher hazard of TB compared to married individuals (aHR = 1.20; 95% CI: 0.71–1.51; *p* = 0.0345). Regarding geographical zones, significantly lower hazards of TB were observed in the Eastern (aHR = 0.21; 95% CI: 0.11–0.42; *p* < 0.001), Western (aHR = 0.37; 95% CI: 0.19–0.73; *p* = 0.004), and Lake zones (aHR = 0.78; 95% CI: 0.18–0.50; *p* < 0.001), compared to the Northern zone. Attending public health facilities remained strongly associated with an increased hazard of TB compared to private/FBO facilities (aHR = 3.92; 95% CI: 2.44–6.28; *p* = 0.001). Similarly, attending health centres (aHR = 0.57; 95% CI: 0.40–0.81; *p* = 0.002) and hospitals (aHR = 0.20; 95% CI: 0.11–0.39; *p* = 0.001) was associated with a reduced hazard of TB compared to dispensaries. Participants in WHO Stage IV had a slightly higher hazard of TB compared to those in Stage I (aHR = 1.01; 95% CI: 1.04–2.31; *p* = 0.042). Breastfeeding individuals also had a higher hazard of TB compared to non-breastfeeding individuals (aHR = 1.12; 95% CI: 1.02–1.56; *p* = 0.034). However, although poor adherence remained associated with an increased hazard of TB, the estimate was imprecise after adjustment (aHR = 1.31; 95% CI: 0.48–2.45; *p* = 0.043), see **Table 4**.

## DISCUSSION

This study aimed at determining median disease-free survival time to diagnosis of TB, incidence of tuberculosis, and determinants associated with diagnosis of tuberculosis among adult PLHIV on ART after IPT completion from 2019 to 2023.

Our study reported the incidence of TB post Isoniazid Preventive Therapy (IPT) as 1.4 per 1000 person-months, which was lower compared to the studies conducted in Uganda [10] with 18.9 per 100,000-person months and Northern Tanzania [11] reported cv 2.08 per 1,000 person-years from 2012 to 2017. Differences may be due to Isoniazid effectively preventing TB in high-risk groups like PLHIV. Early HIV diagnosis and effective ART help maintain stronger immunity, reducing TB risk. Regular TB screening and monitoring enable early detection and treatment before active disease develops.

Also, the study found that 99.3 % of PLHIV had good adherence, which was determined by client self-reporting and viral suppression less than 1000 copies/mil to both ART and IPT regimens that could contribute to better health outcomes and lower incidence of TB [12][13]. Integrated health programs in Tanzania, including those run by NASHCOP and ICAP, focus on both HIV and TB, ensuring coordinated care and better management of co-infections, which could also be another reason contributing to lower tuberculosis incidence [14][15].

The results of this study showed that the overall median disease-free survival time to diagnosis of tuberculosis after IPT completion was 10.12 months, and the duration of diseases free for males was 10.0 months, and female were 10.1 and 8.3 months, respectively. These findings were similar to the studies conducted in Thailand [16], which reported 6 months. Brazil, Rio de Janeiro [17], with 6 months, and South Africa [18], which reported 6-12 months. Furthermore, the findings were similar to those found in South Africa [18] and were lower compared to the 18 months stated in the national guideline for management of HIV and AIDS, 7^th^ edition of 2019, and 2.5 years stated in the Stop TB strategy [19]. This could be due to the emergence of drug resistance, as the development of resistance to isoniazid (INH) can reduce the efficacy of IPT [20]. This resistance may be due to improper use of the medication, such as non-adherence to the prescribed regimen [20]. But also, people living with HIV have an increased risk of TB reactivation and reinfection, which can diminish the protective effects of IPT. The immunosuppressive nature of HIV makes it harder for the body to maintain prolonged protection against TB [20].

In regions with a high TB prevalence, the likelihood of reinfection after completing IPT is significant. This can shorten the duration of protection provided by IPT [18]. Incomplete or irregular adherence to the IPT regimen can lead to suboptimal protection and increase the risk of TB infection or reactivation. Poor nutritional status and co-existing health conditions can weaken the immune system, reducing the effectiveness and duration of IPT [18]. The standard duration of IPT may not be enough to provide long-term protection, especially in high-risk populations. Extended or repeated courses might be necessary, but are not always implemented [18].

The study found that female PLHIV had a higher hazard of tuberculosis compared to male PLHIV. This finding was consistent with the findings in Kenya [21]. This finding can be explained by several factors, such as women experiencing different patterns of HIV disease progression or having different levels of adherence to antiretroviral therapy (ART) and IPT, and also Differences in viral load suppression and CD4 counts could influence the diagnosis of TB [22][23].

Our study also found that clients with lower BMI had a higher hazard of tuberculosis compared to those with normal BMI. This result was in line with the study conducted in Uganda [10] and Eritrea [24]. PLHIV with low BMI might have insufficient nutritional reserves, making them more susceptible to TB even after completing IPT [25]. Also, while IPT is effective in reducing the risk of TB, its effectiveness can be compromised if the individual’s immune system is severely weakened due to low BMI and malnutrition and therefore a stronger immune system supported by good nutrition is crucial for the effectiveness of IPT [25].

Unmarried women exhibited a higher hazard of developing TB compared to married women. This finding is consistent with studies conducted in Kenya [18], Ethiopia [21], and South Africa [15]. These could be explained as married women may benefit from greater economic stability and social support than their unmarried counterparts [22–23]. Such stability can enhance access to healthcare services, adequate nutrition, and improved living conditions, all of which are essential for maintaining a strong immune system and reducing the risk of TB [23]. Also, marital support may positively influence treatment behaviors. Married women may demonstrate better adherence to antiretroviral therapy (ART) and isoniazid preventive therapy (IPT) due to encouragement and support from their spouses [24,25]. Adherence to these regimens is critical in preventing TB among people living with HIV (PLHIV). Additionally, unmarried women may be more vulnerable to psychosocial challenges, including stress, depression, and social isolation, which can adversely affect immune function and increase susceptibility to TB [25,26]. Psychosocial well-being plays a crucial role in overall health and immune competence [26]

The study also found that PLHIV who completed Isoniazid Preventive Therapy (IPT) and were breastfeeding had a higher hazard of tuberculosis (TB), which can be explained in terms of how IPT protection might be affected during breastfeeding.

Our study also found that PLHIV who completed IPT and attended health centers and hospitals had a lower hazard of tuberculosis (TB) compared to those who attended dispensaries. This finding is consistent with the study conducted in China [26]. This could be explained by several reasons, such as health centers and hospitals usually having more advanced diagnostic tools and laboratory facilities, which allow for early detection of TB, HIV-related complications, and other infections, enabling timely interventions that can prevent TB even after IPT completion[26]. Also, they are better equipped, have more trained healthcare professionals, and follow more rigorous protocols for monitoring and managing patients, which can enhance the effectiveness of IPT. Hospitals and health centers often provide comprehensive healthcare services, including regular follow-ups, nutritional support, and management of comorbidities [26].

Furthermore, the study found that PLHIV who completed isoniazid preventive therapy (IPT) and attended public health facilities had a higher hazard of tuberculosis (TB) diagnosis compared to those attending private facilities. This finding is consistent with a study conducted in India [27]. Several explanations may account for this observation. Public health facilities may have stronger adherence to national TB diagnostic guidelines, particularly in the use of microbiological testing, which could lead to increased detection and reporting of TB cases [27]. In contrast, private sector providers, while often associated with better client experience, may have lower adherence to standardized diagnostic protocols, potentially resulting in underdiagnosis or delayed diagnosis of TB [27]. Additionally, public facilities often serve a higher volume of patients, including those from lower socioeconomic backgrounds who may present with more advanced disease or higher underlying risk of TB, further contributing to the observed higher hazard.

### Strengths and limitations

The study was conducted among adult PLHIV aged 15 years and above who completed IPT from the 26 regions of Tanzania mainland, which contributes to the adequate sample size appropriate to detect the desired outcome, and the study has adequate statistical power to precisely detect the association between the exposure and outcome variable, as well as TB incidence.

A limitation of this study is that the study design was a retrospective cohort studies are prone to bias and uses secondary data that cannot go beyond the documented factors and a lot of missing information and censoring.

## Conclusion

Among adult PLHIV on ART who completed IPT in mainland Tanzania in this study, the incidence of TB remained low, confirming the protective benefit of IPT. However, the median time to TB development of approximately 10 months suggests that the protective effect of IPT may wane within the first year after completion. TB risk varied across demographic and clinical groups, with higher risk observed among females, unmarried individuals, and those who were breastfeeding, while older age groups, certain geographic zones, and receiving care at higher-level health facilities were associated with reduced risk. These findings highlight the need for targeted post-IPT monitoring and consideration of additional or extended TB preventive strategies for high-risk groups.

The findings of our study show that the median duration of IPT protection was 10.2 months, which is lower compared to the 18 months stated in the Tanzania HIV treatment guideline. We therefore recommend that frequent and intensive TB screening should be conducted for all PLHIV clients 10 months after IPT completion. Based on the findings from this secondary data analysis, females had higher hazards of tuberculosis incidences; therefore, we recommend targeted TB screening for this group, which should be considered immediately after 8 months since IPT completion. To the Tanzania Ministry of Health, we recommend revising the IPT guidelines and formulating a policy to accommodate a new regimen of TB Preventive Therapy in the treatment guideline

## Data Availability

All data produced in the present study are available upon reasonable request to the authors

